# The “throwaways”. Conflicts of interest in dermatology publications

**DOI:** 10.1101/2021.01.17.21249994

**Authors:** Jorge Roman, David J. Elpern

**Affiliations:** The Ronald O. Perelman Department of Dermatology, New York University Langone Health, New York, New York, United States; The Skin Clinic, Williamstown, Massachusetts, United States

**Keywords:** pharmaceutical industry, continuing medical education

## Abstract

**Importance:** Conflict of interest as it relates to medical education is a topic of concern. Dermatology journals, periodicals, editorials, and news magazines are influential resources that are not uniformly regulated and subject to influence from the pharmaceutical industry.

**Objective:** This study evaluates industry payments to physician editorial board members of common dermatology publications, including “throwaway” publications.

**Design:** A list of editorial board members was compiled from a collection of clinical dermatology publications received over a 3-month period. To analyze the nature and extent of industry payments to this cohort, payments data from the Open Payments database from 2013 to 2019 were collected. Analysis of the total payments, number of transactions, categories of payments, payment sources, and physician specific characteristics was performed.

**Results:** Ten publications were evaluated, and payments data for 466 physicians were analyzed. The total compensation across all years was $75,622,369.64. Services other than consulting, consulting, and travel/lodging payments comprised most of the payments. A faction of dermatologists received the majority of payments. The top payers were manufacturers of biologic medications. Payment amounts were higher for throwaway publications compared to peer-reviewed journals.

**Conclusions:** Editorial board members of dermatology publications received substantial payments from the pharmaceutical industry. A minority of physicians receive the lion’s share of payments from industry. “Throwaway” publications have more financial conflict of interest than peer-reviewed journals. The impact of these conflicts of interest on patient care, physicians practice patterns, and patient perception of physicians is noteworthy.

## Introduction

Health care professionals across all specialties utilize a myriad of resources for staying up to date on the medical literature. While peer reviewed journals are touted as the gold standard, little attention has been given to the role of “throwaway” journals in keeping clinicians abreast of advances in the literature. Throwaway journals are characterized as publications that contain no original investigations, are provided free of charge, have a high advertisement-to-text ratio, are non-society publications, and are seldom peer reviewed.^1^ Previous studies have shown that throwaway journals are more widely read than some peer-reviewed journals.^1,2^ Throwaway journals are attractive to practicing clinicians given their ease of readability. The use of color, larger font size, graphics, and short summaries improve the appeal of throwaway journals to their readership.^3^

Industry-physician interaction is common in all medical specialties, and dermatology is no exception.^4^ Previous studies have examined conflicts of interests among authors of dermatology textbooks, dermatology patient advocacy organizations, dermatology clinical practice guideline authors, and clinical trials in dermatology.^5-9^ Under the Physician Payment Sunshine Act, a part of the Affordable Care Act, payments and other transfers of value by manufacturers and group-purchasing organizations to physicians are reported to the Centers for Medicare and Medicaid Services. These payments are reported in categories including consulting, speaking fees, food, travel, and research.^10^

Given the important role that journals play in the education and clinical practice of dermatologists, we sought to characterize the extent and nature of industry payments to editorial board members of different dermatologic publications, including the throwaway journals. Specifically, we examined the number, amount, and type of payments received, the companies that are contributing the payments, and physician-specific characteristics (sex, practice setting, fellowship training).

## Methods

### Study sample

To replicate a real-world scenario, publications related to clinical dermatology received by author J.R. (a dermatology resident) over a 3-month period were collected. All publication types including peer reviewed journals, non-peer reviewed journals, and periodicals such as news magazines and tabloids were included for analysis. A list of editors was compiled by individually reviewing each publication. Editorial board members whose primary affiliation was outside of the United States (U.S.) and non-physicians (i.e. physician assistants and PhDs) were excluded from the study. Editor names were entered into the Open Payments database and all payments data from 2013 to 2019 were collected. Physician specific information on gender, practice setting, and training was collected via examination of professional information and biographies on individual practice websites. This study did not require approval by the Institutional Review Board, as it did not contain human subject research and utilized publicly available data. The STROBE reporting guidelines were used for this study.^11^

### Data analysis

Data analysis was completed using Excel version 16.41. Descriptive statistics such as mean, median, interquartile range (IQR), and percentages were calculated. Median and IQR were used when appropriate as descriptors when there was a skewed distribution. The statistical significance of intergroup differences was tested by using an independent sample t-test. A two-tailed P-value less than 0.05 was considered as statistically significant.

## Results

Ten publications were evaluated, and 466 individual physicians were identified. The publications included 5 periodicals and 5 journals. The group was comprised of 267 (57.3%) men. The proportion of physicians in academic and private practice settings were almost equal with 242 (51.9%) in private practice and 224 (48.1%) in academic settings. However, of those in private practice, 164 (67.8%) also held academic appointments. Ninety-eight (21%) served on more than 1 editorial board. Further physician characteristics are shown in Table 1.

**Table 1.**
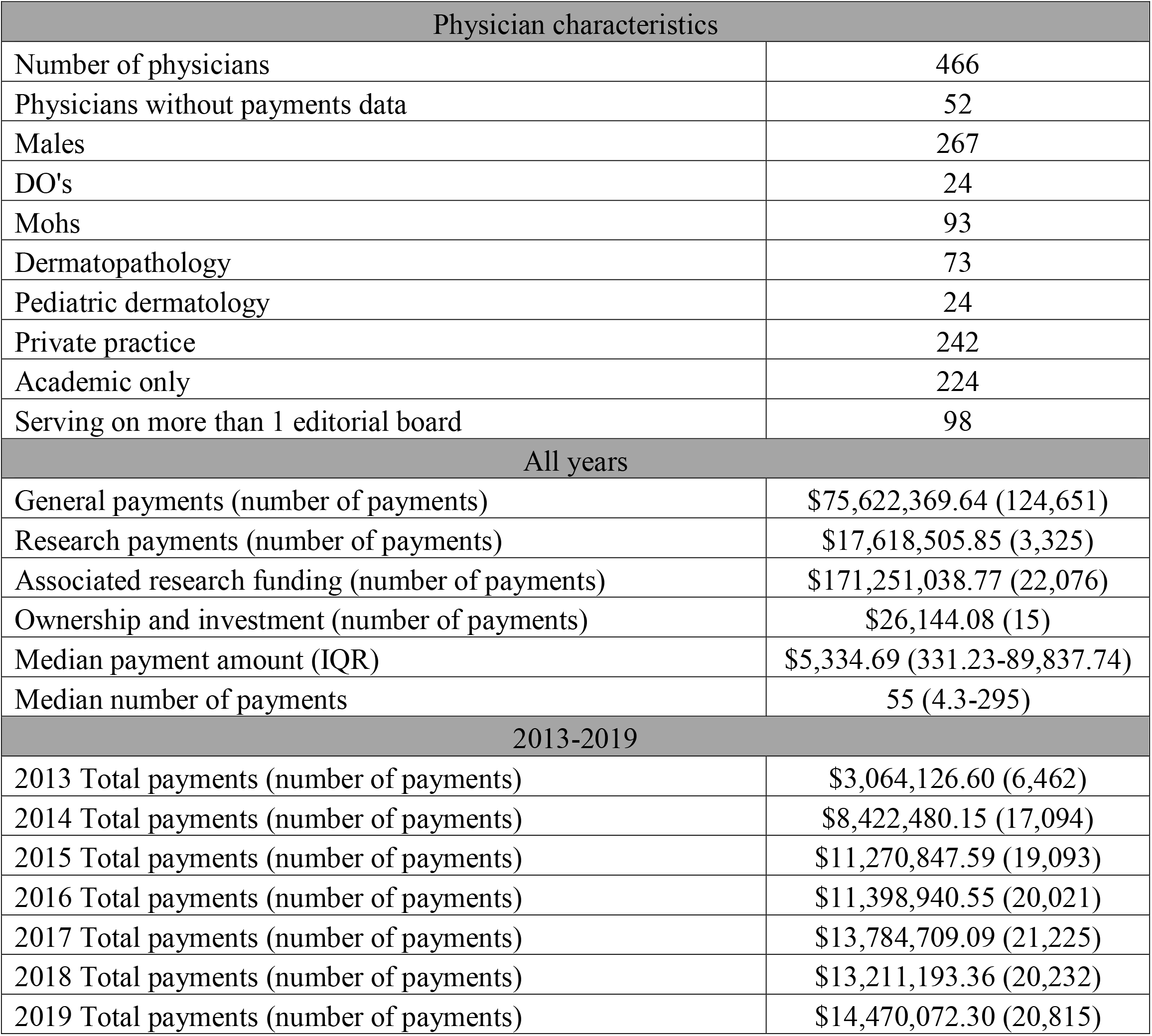
Physician characteristics and overall payments data.

| Physician characteristics |  |
| --- | --- |
| Number of physicians | 466 |
| Physicians without payments data | 52 |
| Males | 267 |
| DO's | 24 |
| Mohs | 93 |
| Dermatopathology | 73 |
| Pediatric dermatology | 24 |
| Private practice | 242 |
| Academic only | 224 |
| Serving on more than 1 editorial board | 98 |
| All years |  |
| General payments (number of payments) | \$75,622,369.64 (124,651) |
| Research payments (number of payments) | \$17,618,505.85 (3,325) |
| Associated research funding (number of payments) | \$171,251,038.77 (22,076) |
| Ownership and investment (number of payments) | \$26,144.08 (15) |
| Median payment amount (IQR) | \$5,334.69 (331.23-89,837.74) |
| Median number of payments | 55 (4.3-295) |
| 2013-2019 |  |
| 2013 Total payments (number of payments) | \$3,064,126.60 (6,462) |
| 2014 Total payments (number of payments) | \$8,422,480.15 (17,094) |
| 2015 Total payments (number of payments) | \$11,270,847.59 (19,093) |
| 2016 Total payments (number of payments) | \$11,398,940.55 (20,021) |
| 2017 Total payments (number of payments) | \$13,784,709.09 (21,225) |
| 2018 Total payments (number of payments) | \$13,211,193.36 (20,232) |
| 2019 Total payments (number of payments) | \$14,470,072.30 (20,815) |

### Overall payments

The total compensation across all years was $75,622,369.64 and the total number of payments was 124,651. Fifty-two (11.2%) physicians had 0 payments reported. The median total industry payment was $5,334.69 (IQR, $331.23-89,837.74). This was higher than the median payment amount averaged across 2013 to 2019 for all U.S dermatologists ($376.37) as well as the median payment for physicians across all specialties ($1083.94).^12^ The median number of payments was 55 (IQR, 4.3-295). This was also higher compared to the median number of payments for all dermatologists and U.S. physicians across all specialties, with medians of 12 and 4 respectively. With the exception of 2017-2018, the total payment and number of payments increased yearly (Table 1).

Of the total payments (total amount), services other than consulting ($31,392,593.02), consulting ($22,201,879.20), and travel/lodging ($8,071,910.76) payments comprised 81.6% of payments (Figure 1, supplementary table 1). Associated research funding and research payments across all years totaled $171,251,038.77 and $17,618,505.85 respectively. It was noted that only a small percentage of the cohort received any kind of payments for associated research funding or research, 33% and 26% respectively. Of those who received payments the median payment amount for associated research funding was $204,284.45 (IQR $39,659.32-960,049.20) and for research payments was $24,484.15 (IQR $5,017.50-144,941.78).

**Figure 1.**
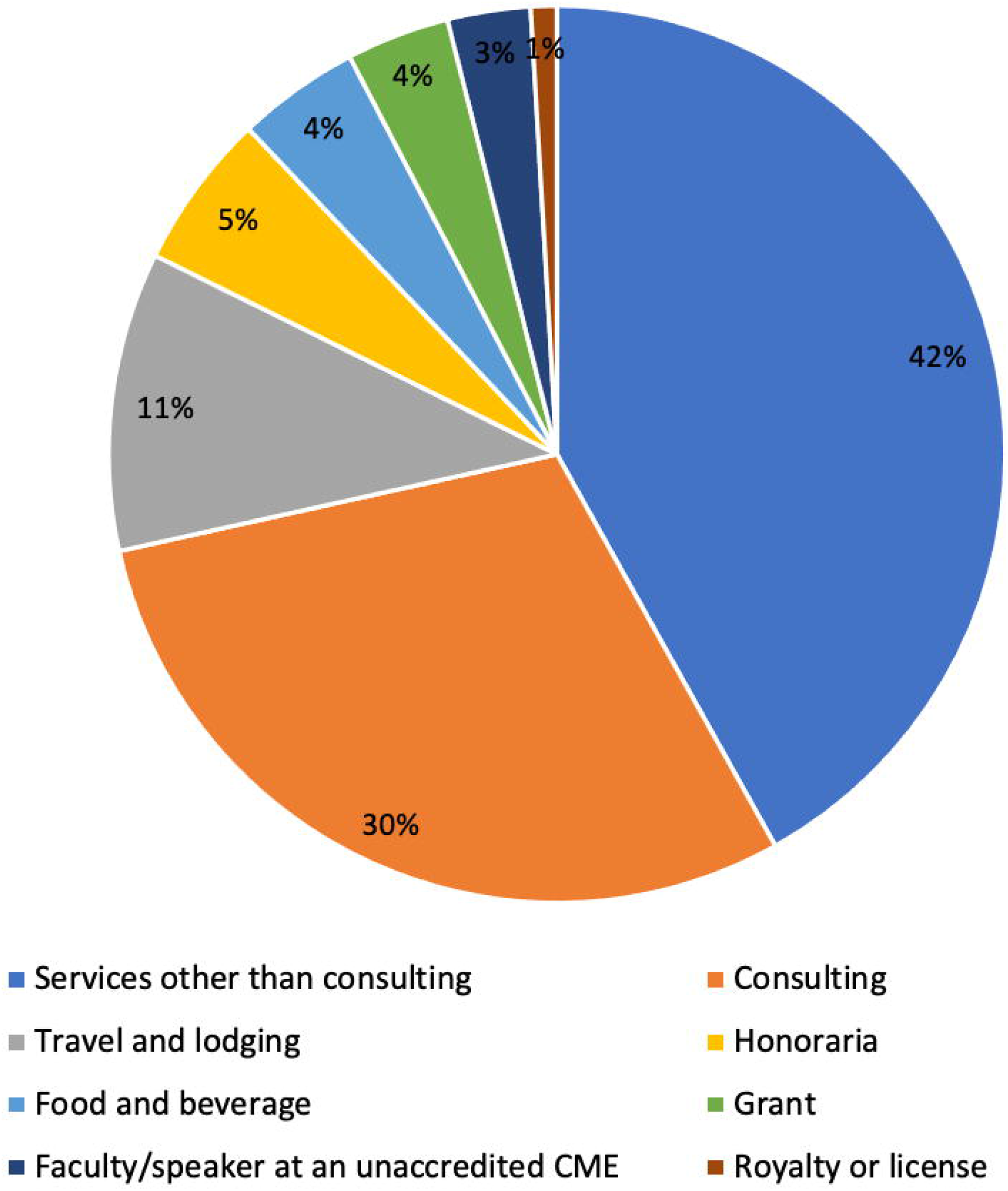
Payments by category.

### Academic vs private

Further analysis was performed after splitting the cohort by practice setting (academic vs private). Compared to those in academic settings, physicians in private practice had higher payments across all categories. The difference in payments was statistically significant for total general payments but not for research payments or associated research funding. Payment differences in the categories of services other than consulting, food and beverage, education, honoraria, and gift were also found to be statistically significant (Table 2).

**Table 2.** Comparison of payments between physicians in academic versus private practice settings.

|  | Academic | Private | P |
| --- | --- | --- | --- |
| Total general payments (number of payments) | \$26,210,268.14 (31,001) | \$49,412,101.50 (93,650) | **0.025** |
| Total research payments (number of payments) | \$5,505,040.83 (1,234) | \$12,113,465.02 (2,091) | 0.09 |
| Associated research funding (number of payments) | \$67,213,288.37 (7,946) | \$104,037,750.40 (14,130) | 0.32 |
| Median for total general payments (IQR) | \$1,048.69 (94.46-19,575.68) | \$19,743.92 (1750.20-18,5248.15) | |
| Median number of total payments (IQR) | 11 (1-63) | 173 (41-463) |  |
|  | Categories of payments |  |  |
| Consulting (number of payments) | \$9,601,983.18 (2,981) | \$12,599,896.02 (3,838) | 0.44 |
| Services other than consulting (number of payments) | \$9,986,340.41 (3,545) | \$21,406,252.61 (7,810) | **0.019** |
| Travel and lodging (number of payments) | \$3,338,185.31 (8,634) | \$4,733,725.45 (13,608) | 0.31 |
| Food and beverage (number of payments) | \$807,956.04 (14,601) | \$2,505,852.32 (63,696) | **<0.001** |
| Education (number of payments) | \$66,933.34 (529) | \$192,825.57 (2,969) | **0.01** |
| Current or prospective ownership or investment interest (number of payments) | \$84,830.47 (4) | \$256,278.64 (2) | 0.52 |
| Honoraria (number of payments) | \$1,036,814.31 (288) | \$3,172,273.45 (829) | **0.02** |
| Faculty/speaker at an unaccredited/non-certified CME (number of payments) | \$777,345.36 (268) | \$1,463,257.45 (576) | 0.18 |
| Gift (number of payments) | \$2,161.24 (126) | \$73,895.71 (401) | **0.02** |
| Grant (number of payments) | \$482,568.48 (18) | \$2,306,195.79 (58) | 0.12 |
| Faculty/speaker at an accredited CME (number of payments) | \$14,150 (6) | \$6,310.50 (6) | 0.41 |
| Entertainment (number of payments) | \$0 (0) | \$101.85 (2) | 0.16 |
| Royalty or license (number of payments) | \$11,000 (1) | \$690,334.87 (4) | 0.32 |

### Top earners

The top 10% of physicians receiving payments collectively received $56,060,893.28 which represented 74.1% of the total payment amount for the entire study group. Seventy-seven percent of this subgroup received payments for research and associated research funding. In total this cohort received $102,076,943.74 in associated research funding and $9,348,517.09 in research payments across all years, accounting for 59.6% and 53.1% of all payments in those categories respectively. This group was comprised of mostly men (78.3%) and the majority (71.7%) work in private practice. Of those in private practice, 78.8% also hold academic appointments. Twenty-five physicians served on more than one editorial board (2.96 on average) (Table 3).

**Table 3.** Characteristics of the top 10% of physicians receiving payments.

| Top 10% |  |
| --- | --- |
| Number of physicians | 46 |
| Total general payments (number of payments) | \$56,060,893.28 (65,644) |
| Total research payments (number of payments) | \$9,348,517.09 (1,832) |
| Total associated research funding (number of payments) | \$102,076,943.74 (12,715) |
| Males | 36 |
| DO's | 2 |
| Mohs | 5 |
| Dermatopathology | 3 |
| Pediatric dermatology | 3 |
| Private practice | 33 |
| Academic only | 13 |
| Serving on more than 1 editorial board | 25 |

### Top payers

The top 20 companies making payments were pharmaceutical manufacturers and combined, paid $64,774,389.91, representing 85.7% of total disbursement. The majority of the companies are manufacturers of biologic medications (Table 4).

**Table 4.**
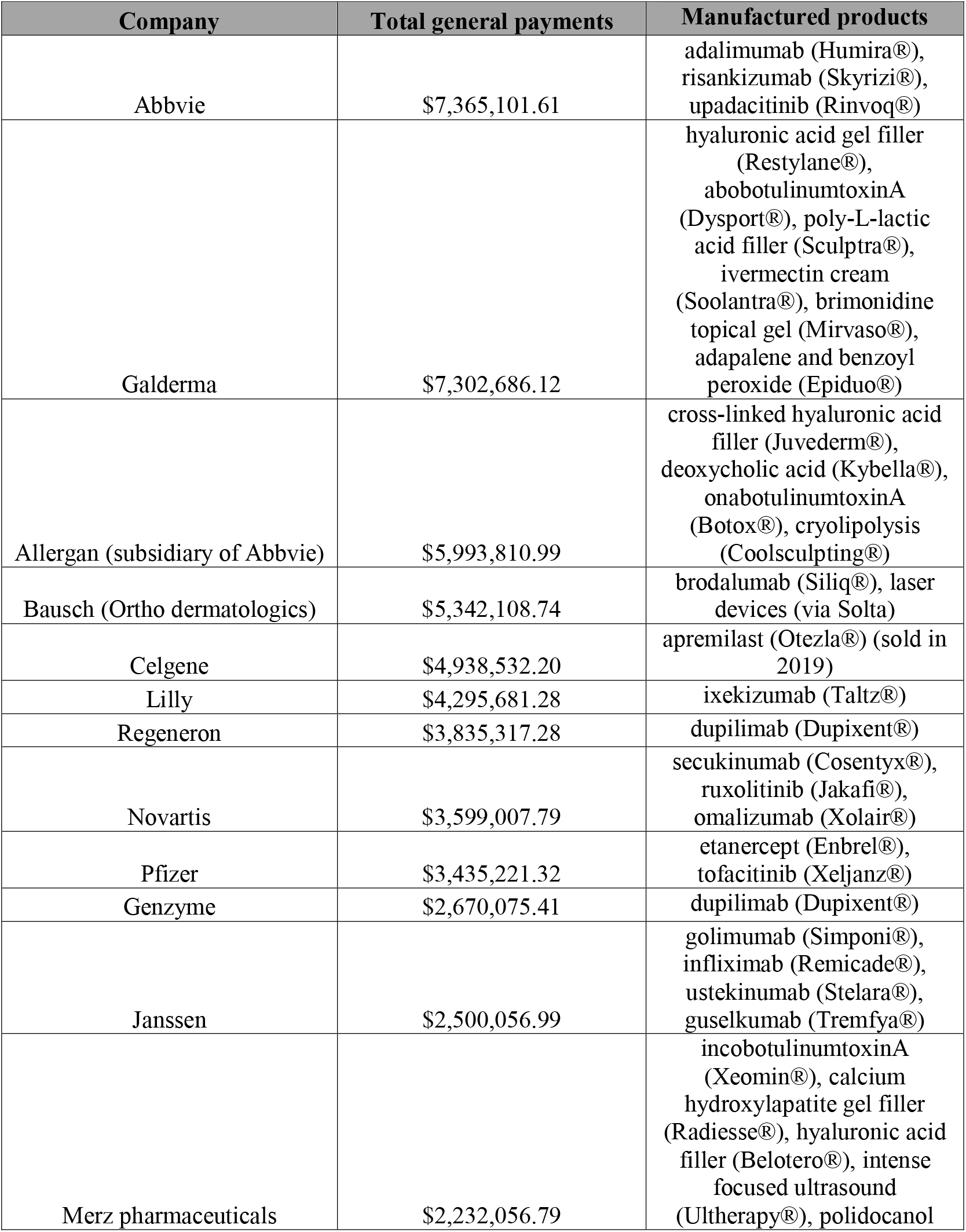
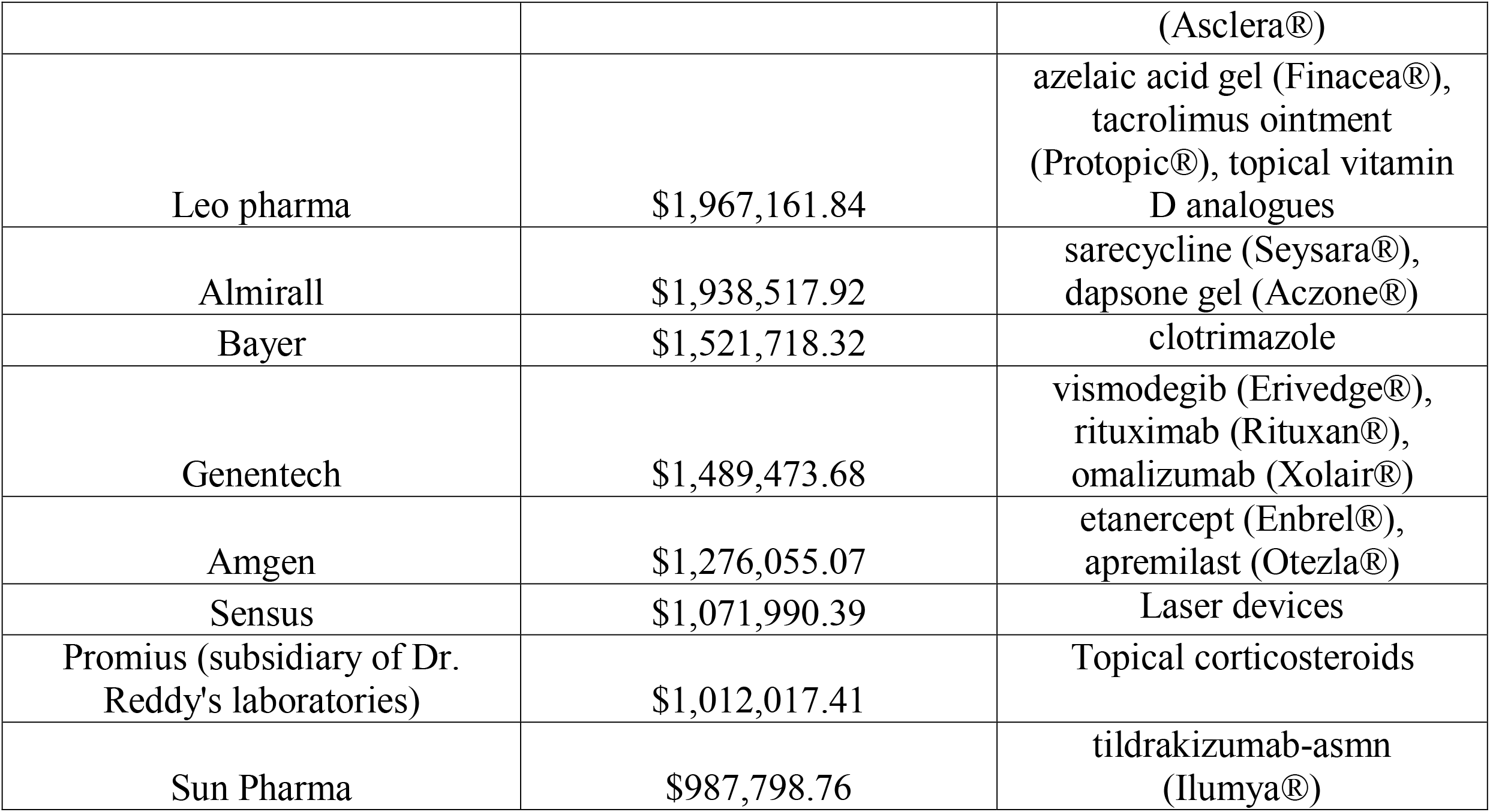
Highest paying companies.

### Individual journal analysis

Payments data for each individual publication was also performed. For simplicity the publications were categorized into 2 groups, periodicals (including news magazines, tabloids, and editorials) and peer-reviewed journals. The average number of editorial board members for periodicals (26.2) was lower than the average for peer-reviewed publications (97.4). The averaged median payment amount ($113,877.02) to physicians on the editorial board members of the periodical publications was 3.5 times higher than to those on editorial boards of peer-reviewed publications ($32,670.59). Associations with professional societies, patient advocacy organizations, access requirements, and other journal data are shown in Supplementary table 2.

## Discussion

In this study we characterized payments from industry to editorial board members of clinical dermatology publications used as important resources in dermatology education and clinical practice. Our study shows that members of editorial boards of various types of publications have ties to industry. Exploration of this group demonstrates a facet of the medical industrial complex that pervades medicine. The data from this study showed that the remuneration received by editorial board members was on average 14 times higher compared to dermatologists at large. Compensation for speaker fees, consulting, travel, and lodging made up the majority of the total payments. The 20 highest-paying manufacturers and most of the companies making payments to dermatologists belong to the pharmaceutical industry. Dermatology as a specialty is a valued target for the pharmaceutical industry, being a relatively small field that treats a number of common and chronic conditions. Dermatologists are one of a few specialties that prescribe high dollar biological medications. Eleven of the top 20 paying companies in our study are manufacturers of biologic medications. Biologics for the treatment of psoriasis is a multibillion-dollar industry and are some of the top-grossing medications in the world. Adalimumab (Humira®) has been the top selling drug for several years with over 19 billion dollars in global sales in 2019 alone.^13^ Since gaining Food and Drug Administration approval for the treatment of adults with moderate to severe atopic dermatitis, dupilimab (Dupixent®) sales have skyrocketed into the billions. The predominance of pharmaceutical payments in dermatology differs from other specialties such as orthopedic surgery, otolaryngology, and ophthalmology, in which device and diagnostic companies provide a greater amount of support.^14-16^

Over the last several decades the number of mediums by which clinicians acquire information to stay abreast of changes in their respective fields have increased. Historically peer-reviewed journals were the mainstay source of information. With advancing technology, the market for resources that clinicians use for continuing medical education (CME) has expanded dramatically to embrace new formats such as podcasts, webinars, virtual conferences, and social media applications (Instagram, Facebook, Twitter). While most academic organizations have guidelines and policies to minimize or prevent conflicts of interests in publishing or the dissemination of information, the same cannot be said for other avenues that are independently sponsored or promoted. Today, most physicians when faced with a surplus of journals do not have the time to critically appraise each individual article. Instead of trying to grasp increasingly complex science or statistical methods, the more practical thing is to read summaries or condensations of educational material. This has paved the way for the “throwaway” journals. Throwaway journals usually do not contain any original investigations, have a high advertisement to text ratio, and are often provided free of charge as they are funded by pharmaceutical companies. These journals are seldom peer-reviewed, but quite popular given their ease of readability.^10^ The articles are frequently written by employees of the publication’s sponsoring companies and are seldom fact-checked by independent reviewers. They often include capsule summaries of conferences, journal article synopses, or therapeutic guides; all sandwiched between pharmaceutical ads. The information within throwaway journals is rarely antagonistic towards the pharmaceutical companies funding the journal.^17^ Despite the popularity of throwaway journals and their direct role in clinician education, there is a dearth of research or discussion of throwaways in the literature. A PubMed search for throwaway journals returns just 8 results, the last article being published in 2005.^18^

The distinction between a classic throwaway and a prototypical peer-reviewed journal has become increasingly difficult. For example, one of the periodicals (DermWorld) that was examined in this study was affiliated with a peer-reviewed journal (Journal of the American Academy of Dermatology (JAAD)). The median payment amounts ($693.68, $1885.65) for both of these publications were the lowest (DermWorld) and second lowest (JAAD) in each of their groups respectively. In contrast, the median payment amount for one of the peer-reviewed journals ($146,159.48, Journal of Clinical and Aesthetic Dermatology (JCAD)) was 11 times greater than the next highest median payment for peer-reviewed journals ($12,526.52, Journal of Drugs in Dermatology). JCAD is a peer-reviewed, PubMed-indexed publication platform wherein every article published is available as full-text and free via PubMed, but with costs covered by advertising and subscriptions. With the commercialization of medical literature and a move toward open access type publishing, these hybrid types of journals are increasingly common. The bias is clear, and the conflicts of interest run deep. Affiliations with other entities including patient advocacy organizations, specialty societies, and other organizations that also receive financial support from the pharmaceutical industry adds another layer of complexity to the relationship.

It stands to reason that industry companies would select for well-known authorities and leaders in the field to provide their knowledge and expertise when evaluating their products. Historically, key opinion leaders (KOLs) earned their positions by performing original research, discovering new therapies, and advancing the field. KOLs are well-known in their respective fields, recognized as the authors of innovative journal articles, senior editors of major textbooks, specialty committee or leadership members, clinical practice guidelines authors, expert speakers at societal meetings, and institutional faculty leaders. Traditionally the road to becoming a KOL involved years of research, teaching, and dissertation. However currently some have asserted that becoming a KOL is more of a commercial enterprise carried out by the pharmaceutical industry and private KOL consulting firms.^19,20^ A usual and effective method for industry to disseminate information is through peer advocacy.^21^ This practice makes sense from a business standpoint as KOLs are valuable figures that can lend expertise and credibility to new pharmaceuticals. Depending on the need, whether a company is looking to introduce a new product, re-brand a previous or newly reformulated product, or develop CME programs, KOLs can function as medical brand ambassadors to target specific audiences. The marketing value of KOLs is analogous to celebrity sponsorship deals in commercial ventures. The line between a trusted colleague sharing their knowledge and a salesperson selling a product is consequently blurred. In an unadulterated world, delivery of information by KOLs would be moral if the material was impartial and rooted in evidence-based medicine. However complete objectivity seems questionable when one party benefits so greatly. Industry offers many advantages to KOLs including paid consultancy, participation in clinical trials, prestige in the eyes of peers, and opportunities for article authorship. The medical literature represents a useful avenue for industry to take advantage of the credibility and standing of KOLs.^22^ The web of interaction is broad as evidenced by the activities of the top paid dermatologists in our study. Many of the top earners serve on multiple editorial boards, hold dual private and academic appointments, and run a conglomerate of CME activities backed by industry with the purpose of influencing dermatologists at large. As examples, the highest earner received payments from 53 different companies and one physician in the top 10% serves on 6 editorial boards, including several of the peer-reviewed journals. Eighty-eight percent of physicians in this study received payments from industry. This was higher than the percentage reported for dermatology textbook authors (54%) and the percentages reported in the studies by Feng et al. and Checketts et al., 73.3% and 86% respectively.^4,6,23^

Historically, collaboration between physicians and the pharmaceutical industry has resulted in innovations and advancements in medicine. When conducted properly, the relationship between physicians and industry serves to advance the field of medicine as a whole with the ultimate goal of the lives of patients. However, the interests and commitments of physicians should deviate from those of industry. While caring for patients is the primary responsibility of physicians, industry is chiefly concerned with the responsibility to their shareholders. As with any other business, the objectives of industry are geared towards profit. Industry engagement occurs so often that the practice has become a normalized component of physician education. This element of medical education has evolved over several decades and is so ubiquitous that many trainees and clinicians have become anesthetized to the practice. The fraternity of medicine is one in which new inductees observe their teachers and mentors giving industry sponsored lectures, serve on industry advisory boards, and receiving industry funding for research.^24^ These practices are so ingrained in our profession that participation is actually desirable for advancing academic careers or enhancing prestige. The “supportive” role of industry in medical education is ethically problematic.

Patients expect physicians to deliver effective, safe, and compassionate care based on evidence and best practices. As medicine is always changing, physicians must stay abreast of new therapeutics, devices, skills, and treatments. Establishing and upholding standards of competence is a responsibility of physicians to society. When these standards are perverted by industry, patients become unknowing victims of commerce. Over recent years industry has played an increasingly direct role in physician education. The pharmaceutical industry’s exploitation of medicine is alive and well, flourishing through academic literature, commercial marketing, and compliant colleagues. Industry has become so intertwined with medicine that it shapes medical knowledge and opinion to suit its commercial needs. It has injected its presence into clinics, conferences, research, journals, and medical education. This relationship is not completely clandestine. Funding from industry supports research grants, clinical trials, and educational programs. As physicians we need to be aware of how industry influences the information we need to care for our patients. Industry promotion and marketing sways the independence of information presented to clinicians to suit their needs. The quality and integrity of clinician education is paramount in maintaining the public’s trust in our profession. In order to maintain the standards of postgraduate professional education the relationship between industry and accredited education must be made transparent.

## Data Availability

The data is available on a public website.

https://openpaymentsdata.cms.gov/

## Acknowledgements

None

## Supplementary material

**Table 1.** Categories of payments.

| Categories of payments |  |
| --- | --- |
| Services other than consulting (number of payments) | \$31,392,593.02 (11,355) |
| Consulting (number of payments) | \$22,201,879.20 (6,819) |
| Travel and lodging (number of payments) | \$8,071,910.76 (22,242) |
| Honoraria (number of payments) | \$4,209,087.76 (1,117) |
| Food and beverage (number of payments) | \$3,313,808.36 (78,297) |
| Grant (number of payments) | \$2,788,764.27 (76) |
| Faculty/speaker at an unaccredited/non-certified CME (number of payments) | \$2,240,602.81 (844) |
| Royalty or license (number of payments) | \$701,334.87 (5) |
| Current or prospective ownership or investment interest (number of payments) | \$341,109.11 (6) |
| Education (number of payments) | \$259,758.91 (3,498) |
| Gift (number of payments) | \$76,056.95 (527) |
| Faculty/speaker at an accredited CME (number of payments) | \$20,460.50 (12) |
| Entertainment (number of payments) | \$101.85 (2) |

**Table 2.** Individual journal characteristics and payments data.

| Journal characteristics |  |
| --- | --- |
| **Dermatology News** |  |
| Category | Periodical |
| Access | Free |
| Number of editorial board members | 23 |
| Members serving on more than 1 editorial board (%) | 8 (34.8) |
| Total general payments (number of payments) | \$4,459,080.96 (7,707) |
| Total research payments (number of payments) | \$1,007,120.56 (319) |
| Total associated research funding (number of payments) | 18,916,442.59 (2,114) |
| Median total general payments (number of payments) | \$41,326.01 (187) |
| **The Dermatologist** |  |
| Category | Periodical |
| Access | Free |
| Associations | National Psoriasis Foundation, The National Eczema Association, The National Rosacea Society, The Skin Cancer Foundation |
| Number of editorial board members | 32 |
| Members serving on more than 1 editorial board (%) | 25 (78.1) |
| Total general payments (number of payments) | \$20,393,699.41 (23,285.00) |
| Total research payments (number of payments) | \$6,170,311.33 (958) |
| Total associated research funding (number of payments) | \$35,322,265.41 (5,157) |
| Median total general payments (number of payments) | \$ 198,453.28 (383) |
| **Practical Dermatology** |  |
| Category | Periodical |
| Access | Free |
| Number of editorial board members | 47 |
| Members serving on more than 1 editorial board (%) | 28 (59.6) |
| Total general payments (number of payments) | \$23,157,228.57 (33,383) |
| Total research payments (number of payments) | \$4,899,282.62 (687) |
| Total associated research funding (number of payments) | \$33,955,728.77 (3,880) |
| Median total general payments (number of payments) | \$244,372.97 (428) |
| **Dermatology Times** |  |
| Category | Periodical |
| Access | Free |
| Number of editorial board members | 12 |
| Members serving on more than 1 editorial board (%) | 9 (75) |
| Total general payments (number of payments) | \$3,660,767.34 (6,348.00) |
| Total research payments (number of payments) | \$2,155,154.69 (462) |
| Total associated research funding (number of payments) | \$18,325,820.76 (2,546) |
| Median total general payments (number of payments) | \$84,539.15 (380) |
| **DermWorld** |  |
| Category | Periodical (affiliated with peer-reviewed journal) |
| Access | Paid subscription |
| Associations | American Academy of Dermatology |
| Number of editorial board members | 17 |
| Members serving on more than 1 editorial board (%) | 5 (29.4) |
| Total general payments (number of payments) | \$284,468.87 (1,100) |
| Total research payments (number of payments) | \$72 (1) |
| Total associated research funding (number of payments) | \$115,985.54 (49) |
| Median total general payments (number of payments) | \$693.68 (5) |
| **Cutis** |  |
| Category | Journal reviewed by editorial board members, indexed in MEDLINE® |
| Access | Free |
| Associations | Association of Military Dermatologists, Skin of Color Society, Society of Dermatology Hospitalists |
| Number of editorial board members | 114 |
| Members serving on more than 1 editorial board (%) | 40 (35.1) |
| Total general payments (number of payments) | \$18,590,790.23 (26,960) |
| Total research payments (number of payments) | \$5,872,727.48 (1,164) |
| Total associated research funding (number of payments) | \$43,050,936.87 (4,399) |
| Median total general payments (number of payments) | \$2,628.25 (37) |
| **JAMA Dermatology** |  |
| Category | Peer-reviewed journal indexed with MEDLINE® |
| Access | Paid subscription |
| Number of editorial board members | 25 |
| Members serving on more than 1 editorial board (%) | 7 (28) |
| Total general payments (number of payments) | \$1,712,629.65 (2,366) |
| Total research payments (number of payments) | \$115,610.91 (111) |
| Total associated research funding (number of payments) | \$3,433,872.97 (548) |
| Median total general payments (number of payments) | \$153.04 (2) |
| **Journal of the American Academy of Dermatology** |  |
| Category | Peer-reviewed journal indexed with MEDLINE® |
| Access | Paid subscription |
| Number of editorial board members | 127 |
| Members serving on more than 1 editorial board (%) | 35 (27.6) |
| Total general payments (number of payments) | \$15,435,274.85 (22,313) |
| Total research payments (number of payments) | \$3,275,731.90 (794) |
| Total associated research funding (number of payments) | \$36,199,238.01 (5,599) |
| Median total general payments (number of payments) | \$1,885.65 (18) |
| **Journal of Clinical and Aesthetic Dermatology** |  |
| Category | Peer-reviewed journal indexed with PubMed but not indexed with MEDLINE® |
| Access | Free |
| Associations | American Cutaneous Oncology Society, The Dermatologic & Aesthetic |
|  | Surgery International League,<br>American Acne & Rosacea Society |
| Number of editorial board members | 91 |
| Members serving on more than 1 editorial board (%) | 52 (57.1) |
| Total general payments (number of payments) | \$39,619,853.99 (60,310) |
| Total research payments (number of payments) | \$12,024,264.82 (1,809) |
| Total associated research funding (number of payments) | \$84,388,373.29 (11,295) |
| Median total general payments (number of payments) | \$146,159.48 (412) |
| **Journal of Drugs in Dermatology** |  |
| Category | Peer-reviewed journal indexed with<br>PubMed and MEDLINE® |
| Access | Free for residents and fellows,<br>complimentary 3-year subscription for<br>dermatologists |
| Associations | International Society of Dermatologic<br>Surgery, Skin of Color Seminar Series,<br>Orlando Dermatology Aesthetic and<br>Clinical Conference |
| Number of editorial board members | 130 |
| Members serving on more than 1 editorial board (%) | 41 (31.5) |
| Total general payments (number of payments) | \$25,071,730.66 (37,238) |
| Total research payments (number of payments) | \$9,172,439.26 (1,272) |
| Total associated research funding (number of payments) | \$55,232,215.29 (6,874) |
| Median total general payments (number of payments) | \$12,526.52 (105) |

